# ACUTE VISION LOSS IN A PATIENT WITH COVID-19

**DOI:** 10.1101/2020.06.03.20112540

**Authors:** Vijairam Selvaraj, Daniel Sacchetti, Arkadiy Finn, Kwame Dapaah-Afriyie

## Abstract

To date, there have been reports of neurologic manifestations in Covid-19 patients including ischemic strokes, Guillain-Barre Syndrome and anosmia. In this case report, we describe a patient who presented with dysosmia, dysgeusia along with monocular peripheral vision loss after being diagnosed with Covid-19.

## INTRODUCTION

As of May 24, 2020, there are more than 5,200,000 cases of SARS-CoV2 worldwide.^1^ To date, there have been several reports of neurologic manifestations in these patients including ischemic strokes, Guillain-Barre Syndrome and anosmia.^2-4^ Wu et al. described a case series involving 38 patients with Covid-19 where 12 patients had ocular signs of epiphora, conjunctival congestion and chemosis.^5^ We report a patient who presented with dysosmia, dysgeusia along with monocular peripheral vision loss after being diagnosed with Covid-19.

## CASE PRESENTATION

A woman in her 50s with history of hypertension, hyperlipidemia and headaches presented to the hospital with fever, chills and cough one week after she tested positive for SARS-CoV2. She reported acute, painless right eye monocular visual disturbance, described as a white cloud and blurriness involving most of her right eye, sparing the superior nasal aspect. She denied any left eye visual disturbances. She denied any other ocular symptoms such as flashers, floaters, or diplopia. She denied any jaw claudication, scalp tenderness, unintentional weight loss. Other neurological symptoms included dysgeusia, dysosmia, right ear hypoascusis and subjective right hemiparesis. She was not taking any medications at home.

On the day of admission, her neurological exam was remarkable for severe right eye vision loss. She was unable to visualize or count fingers in right temporal field and inferior nasal field. Left eye exam was normal. Relative afferent pupillary defect was absent. The rest of her neurological exam was normal. There was no tenderness to palpation of temporal area. The following day, she reported fifty percent improvement in her vision. Her vision in the far periphery of the right eye was blurry but she was able to count fingers in all fields. Visual acuity was 20/70. Dilated fundoscopic exam was normal. Ocular pressures were normal. There was no evidence of optic disc edema, Hollenhorst plaque, retinal whitening or hemorrhages.

Her laboratory values were normal including CBC, BMP and ESR. Her CRP was 7 and d dimer was 206 ng/ml. LDL was elevated at 131. Initial MRI of the brain without gadolinium did not reveal any intraparenchymal or cranial nerve abnormalities, though was notable for a partially empty sella turcica. MRI of the orbits, face and neck with and without gadolinium revealed no area of abnormal enhancement. The optic nerves, chiasm and optic tracts appeared normal. CTA showed no significant carotid disease. Her vision spontaneously improved during her hospitalization and she was discharged home on aspirin and atorvastatin. She was advised to follow up in the Ophthalmology clinic in one month.

## DISCUSSION

The clinical spectrum of illness due to Covid-19 continues to evolve. Acute vision loss is a medical emergency and can occur over a period of a few seconds or minutes to a few days. Vision may become blurry, cloudy, completely or partially absent, or affected by flashes or floaters. Acute vision loss is usually painless but may also be associated with ocular pain, redness and headache. Most cases of visual loss can be diagnosed by history and physical examination alone.

Common causes of acute vision loss include Central Retinal Artery Occlusion (CRAO), Central Retinal Vein Occlusion (CRVO), Retinal Detachment, Optic Neuropathy and Inflammatory conditions such as Giant Cell Arteritis (GCA). Given normal ESR, CRP and absence of classic symptoms such as jaw claudication and scalp tenderness, GCA was less likely. Given absence of optic disc edema, Nonarteritic Ischemic Optic Neuropathy (NAION) was felt to be less likely. CRVO was unlikely due to the absence of retinal hemorrhages and cotton wool spots on fundoscopic exam.

Given history of peripheral monocular vision loss along with normal neurologic and ophthalmologic exam, transient Branch Retinal Artery Occlusion (BRAO) was the most likely diagnosis in our patient. BRAO stems from occlusion of a branch of the Central Retinal Artery and accounts for approximately 38% of all acute retinal artery occlusions. More than 90 percent of patients with BRAO are above the age of 40 and have vascular risk factors such as hyperlipidemia and hypertension. Most patients with BRAO recover normal vision as observed in our case. Given the MRI evidence of a partially empty sella, idiopathic intracranial hypertension, or pseudotumor cerebri, was also considered a possibility for transient visual loss. She did not undergo lumbar puncture to measure intrathecal pressure as her ocular symptoms had improved and denied any headache symptoms.

Previous strains of coronavirus seem to invade the CNS mostly through the hematogenous route but also can invade through the cribriform plate and the conjunctiva.^5,6^ The pathophysiology in our case is unclear. One of the mechanisms could involve inflammation associated with Covid-19 itself although her CRP and ESR were normal.^7^ Another mechanism could be related to the thromboembolic phenomenon seen in Covid-19 patients although our patient’s d dimer was normal and there were no other hematologic abnormalities. Magro et al. showed that there might be a microvascular injury syndrome mediated by activation of complement pathways and an associated procoagulant state that may be at play in these patients.^8^

Our patient’s symptoms were early in the course of her illness and could be useful in the triage of patients. A thorough neurologic exam is essential in all patients diagnosed with Covid-19. This case illuminates broader spectrum of Covid-19-related symptomatology and emphasizes the need for clinicians to be aware of the various clinical manifestations associated with this infection.

## Data Availability

All the data is included within the manuscript text.

## REFERENCES

1. World Health Organization. Coronavirus Disease (COVID-19) Situation Report −125. Accessed: May 24, 2020 https://www.who.int/emergencies/diseases/novel-coronavirus-2019/situation-reports

2. Oxley TJ, Mocco J, Majidi S, et al. Large-Vessel Stroke as a Presenting Feature of Covid-19 in the Young. N Engl J Med. 2020;382(20):e60. doi:10.1056/NEJMc2009787

3. Toscano G, Palmerini F, Ravaglia S, et al. Guillain-Barré Syndrome Associated with SARS-CoV-2 [published online ahead of print, 2020 Apr 17]. N Engl J Med. 2020; NEJMc2009191. doi: 10.1056/NEJMc2009191

4. Eliezer M, Hautefort C, Hamel A, et al. Sudden and Complete Olfactory Loss Function as a Possible Symptom of COVID-19. JAMA Otolaryngol Head Neck Surg. Published online April 08, 2020. doi:10.1001/jamaoto.2020.0832

5. Wu P, Duan F, Luo C, et al. Characteristics of Ocular Findings of Patients With Coronavirus Disease 2019 (COVID-19) in Hubei Province, China. JAMA Ophthalmol. 2020;138(5):575–578. doi:10.1001/jamaophthalmol.2020.1291

6. Baig AM. Neurological manifestations in COVID-19 caused by SARS-CoV-2. CNS Neurosci Ther. 2020;26(5):499–501. doi:10.1111/cns.13372

7. Zhou F, Yu T, Du R, et al. Clinical course and risk factors for mortality of adult inpatients with COVID-19 in Wuhan, China: a retrospective cohort study. Lancet 2020; 395:1054-1062.

8. Magro C, Mulvey JJ, Berlin D, et al. Complement associated microvascular injury and thrombosis in the pathogenesis of severe COVID-19 infection: a report of five cases [published online ahead of print, 2020 Apr 15]. Transl Res. 2020;S1931-5244(20)30070-0. doi:10.1016/j.trsl.2020.04.007

